# Impact of the national test strategy on the development of the Covid-19 pandemic in Denmark

**DOI:** 10.1101/2021.07.08.21260182

**Authors:** Peter K. Busk, Thomas Birk Kristiansen, Allan Engsig-Karup

## Abstract

During the Covid-19 pandemic, Denmark has pursued a unique mass testing strategy culminating in testing of more than 8,000 citizens per 100,000 inhabitants per day in the Spring 2021. The strategy includes free access to Covid-19 testing and since 2021, compulsory documentation for negative test or vaccination for access to workplace, educational institutions, restaurants, and many other places.

In the present study we analysed the effect of the Danish mass testing strategy throughout relevant stages of the pandemic. Mass testing was found not to have impact on the number of hospitalizations during the pandemic. Furthermore, during the highest level of testing in the spring 2021 the fraction of positive tests increased slightly on comparable days indicating that mass testing at best, did not reduce the prevalence of Covid-19.

The relationship between positives in antigen testing and in PCR testing indicated that many patients are not tested early in their infection where the risk of transmission is highest.

A likely explanation for the lack of impact of mass testing on hospitalizations and infections is that the testing increases risk-behaviour of the tested persons and that a high fraction of false negatives at low Covid-19 prevalence combined with increased risk-behaviour outweighs beneficial effects of mass testing.

## Introduction

The first case of SARS-CoV-2 (Covid-19) infection in Denmark was diagnosed on February 27^th^ 2020 (Statens Serum Institut, 2020). Within a month the Danish authorities decided on an aggressive strategy for Covid-19 diagnostic testing (The Danish Ministry of Health, 2020a). In a few months, the capacity for rt-PCR testing for SARS-CoV-2 was expanded to allow every citizen to be tested for free without prior medical evaluation. Thus, in December 2020, Covid-19 testing in Denmark reached 2,900 tests per day per 100,000 inhabitants (Ritchie et al., 2020). By implementation of rapid antigen tests for mass screening purposes the Danish test capacity was further increased and peaked at 12,167 Covid-19 tests per day per 100,000 inhabitants in May 2021. As part of an reopening plan for Denmark in March, 2021 a negative Covid-19 test, completion of Covid-19 vaccination or past Covid-19 became compulsory for attending education, restaurants, fitness centres etc. (The Danish Ministry of Health, 2021). Moreover, children in elementary school were strongly urged to take biweekly Covid-19 tests.

The Danish Covid-19 strategy is unique as no other country has relied so heavily on mass testing of the general population.

In late 2020, Slovakia tested the entire population for Covid-19 twice within a week. This systematic antigen testing reduced the prevalence of the disease by 70 % compared to no testing (Pavelka et al., 2021). The Danish strategy is based partly on voluntary testing and partly on compulsory testing for participation in educational and recreational activity and for admittance to workplace. As testing is unsystematic, there is a large difference in the frequency of testing of each citizen. Although the number of tests since the outbreak of the pandemic corresponds to an average of 11 tests per citizen, by July 1^st^ 2021 approximately 900,000 thousand persons corresponding to 10 % of the population aged more than five years has not been tested even once for Covid-19 (Statens Serum Institut, 2021a).

Recently, the effect of the Danish test strategy was evaluated by comparing actual test data to an extrapolation of the expected number of positive tests if Denmark had followed a low testing strategy like Norway and Sweden, and to an estimation of the total number of contaminated persons in the country (Pedersen et al., 2021). This bottom-up analysis suggests that testing at the scale implemented in Denmark reduces the contact number by 25 %.

The present study assessed the observable impact of the extensive Covid-19 testing of the Danish population primarily by examining the correlation between the number of tests and the number of hospitalizations in meaningful periods of the Covid-19 pandemic.

## Data and methods

Data for number of tests, number of positives and hospitalizations were downloaded from Statens Serum Institut’s daily report on June 2th 2021 (Statens Serum Institut, 2021a).

Data for vaccinations were downloaded on July 5^th^ 2021 (Statens Serum Institut, 2021b)

Correlations were calculated as Pearson correlation coefficient between the data sets (Bravais, 1844).

## Analysis and results

The number of Covid-19 positives detected is expected to depend strongly on the number of tests performed. However, the prevalence and the ability to test the right persons of interest do also influence on the number of Covid-19 positives that are detected. In contrast, hospitalization depends on the patient’s symptoms and is thus a more suitable measure of the development of the pandemic. Hence, the number of hospitalizations is expected to show a negative correlation to the number of tests if mass testing reduces Covid-19 infections in the population.

Figure 1 shows the number of Covid-19 tests and hospitalizations since the first patient diagnosed with Covid-19 in Denmark was hospitalized on March 1^st^ 2020 until June 21^st^ 2021. The mass testing can be divided into stages 0 - 3:

**Figure 1.**
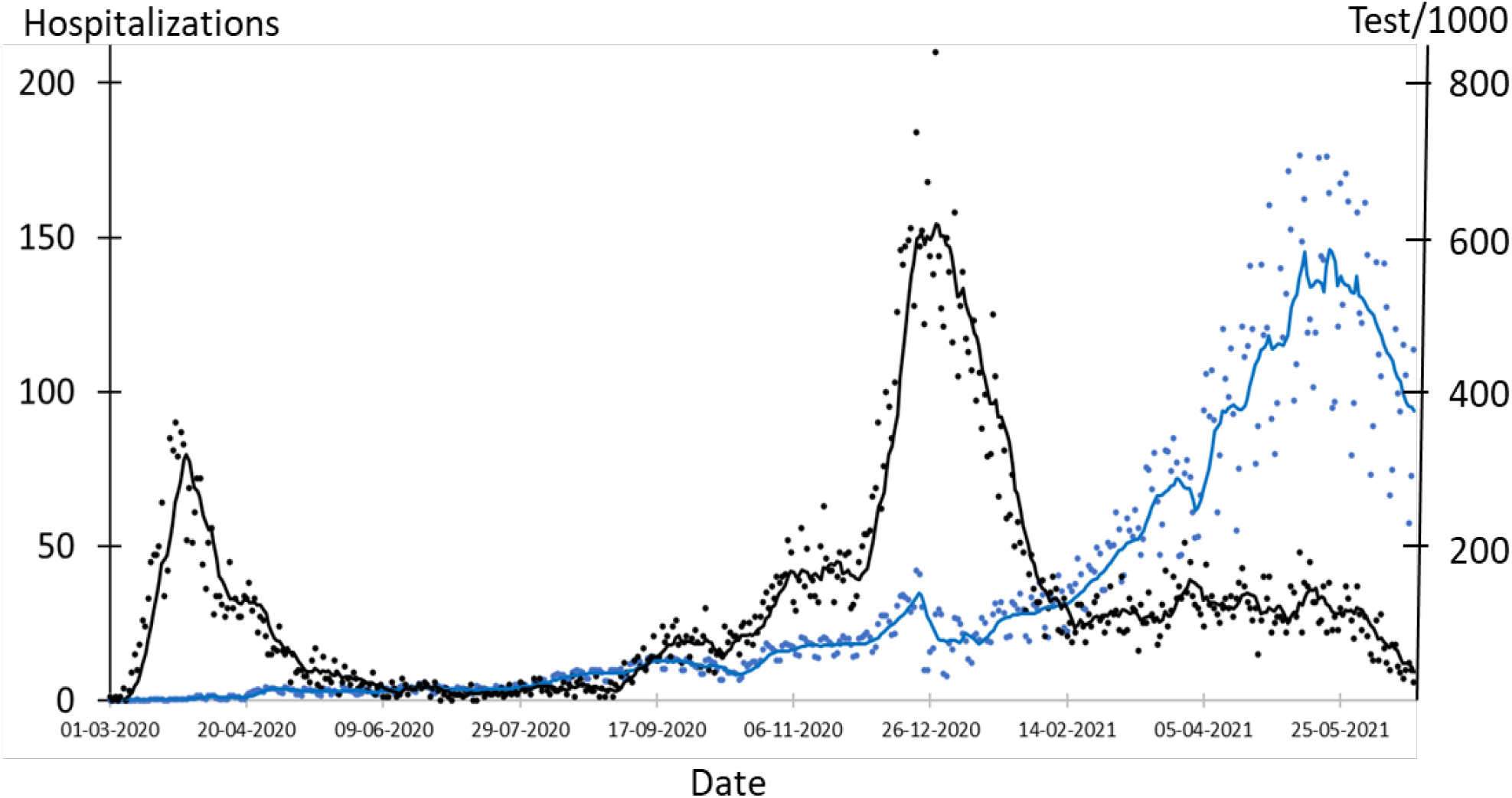
Number of Covid-19 tests and hospitalizations in Denmark during the pandemic. Blue dots: Number of tests. Blue line: Seven days average of number of tests. Black dots: Hospitalizations. Black line: Seven days average of hospitalizations.

0. **March 1^st^ 2020 - May 26^th^ 2020** where test capacity was set up. The number of daily tests increased from 49 to 15,980. This period includes the lock down period March 12^th^ 2020 - May 26^th^ 2020 (The Danish Government, 2020; The Prime Minister’s Office, 2020).
1. **May 27^th^ 2020 – December 6^th^ 2020** when the Danish society was open with relatively few changes in restrictions such as introduction of compulsory facemasks in public transport on August 22^nd^ and all indoor facilities with public assess on October 29^th^ (The Danish Ministry of Health, 2020b, 2020c). During this stage testing increased from 14,781 tests per day to 69,215 tests per day.
2. **December 11^th^ 2020 – February 28^th^ 2021.** The Winter lockdown period with severe restrictions (The Danish Ministry of Health, 2020d). Hospitalizations peaked at 210 on December 28^th^ and testing increased from 102,431 tests per day to 143,001 tests per day. Vaccinations started on December 27^th^ and reached 7.3 % of the population, mostly elderly on February 28^th^ (Danish Health Authority, 2021; Ritchie et al., 2020).
3. **April 6^th^ 2021 – June 21^st^ 2021.** Schools, other educations and liberal professions were opened (Nationalt Kommunikationspartnerskab Covid-19, 2021). Malls etc. were opened on April 21^st^ and many indoor facilities such as restaurants, museums etc. opened on May 6^th^ for guests that had been vaccinated or had a negative Covid-19 test not older than 72 hours (The Danish Government, 2021a). The requirement for facemasks was abandoned on June 14^th^ (The Danish Government, 2021b). This period includes the most extensive level of mass testing with an average of 466,861 tests per day corresponding to 8,049 tests per day per 100,000 persons. By the end of May, all citizens above 65 years had been offered a Covid-19 vaccine and 37.4 % of the population had received the first dose (Danish Health Authority, 2021; Ritchie et al., 2020). On June 21^st^ 53 % of the population had received the first dose.

The incubation period for Covid-19 until a patient is infectious and test positive in an rt-PCR or an antigen test is 3 – 7 days (Cevik et al., 2021; Mallett et al., 2020; World Health Organization, 2020). Without testing and quarantine the index patient (patient 1, Figure 2) can infect the next patient (patient 2, Figure 2). After additional 7 – 13 days patient 2 will be hospitalized (Faes et al., 2020). Thus, there is a considerable delay from the infectious contact between patients 1 and 2 at day x to the hospitalization of patient 2 on day x+7 to x+13 (Figure 2). As the window of passing Covid-19 to others is 3 – 10 days post infection x can vary from 3 to 10 implying that hospitalization of patient 2 happens 10 to 23 days after infection of patient 1.

**Figure 2:**
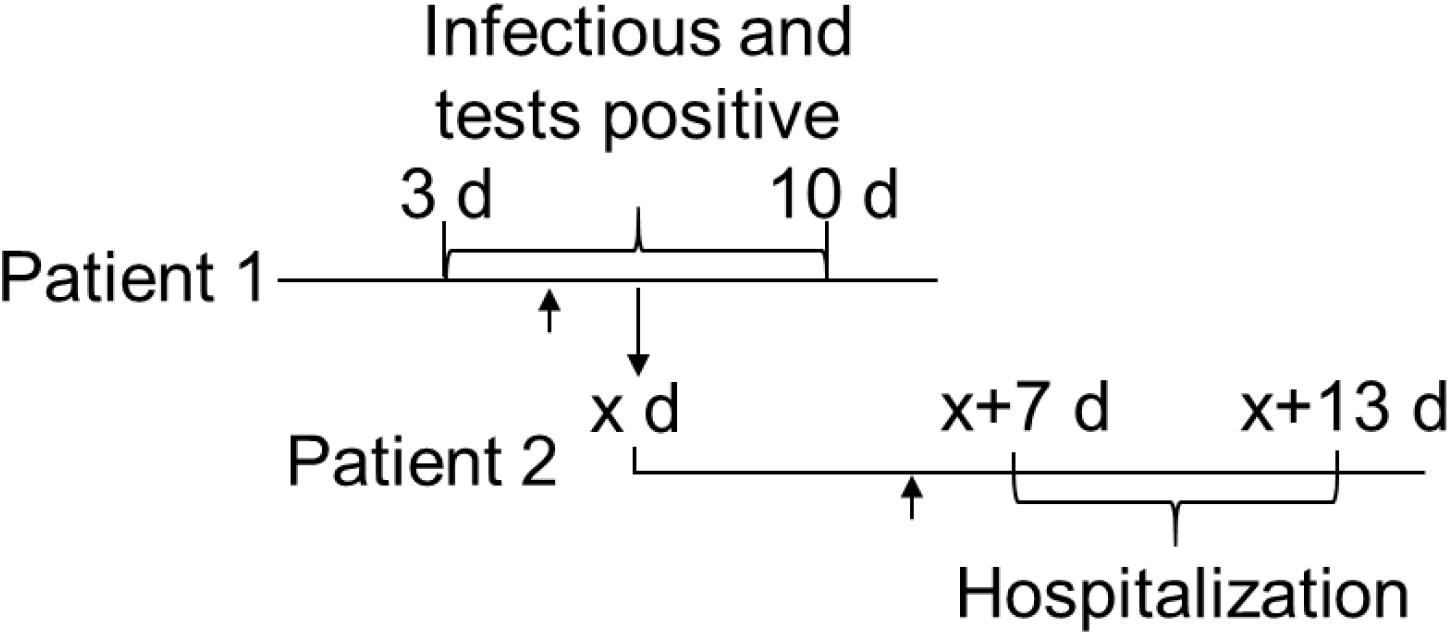
Overview of timing of infectivity, positive test, and hospitalizations for two patients. Patient 1 is infected on day 0 and can infect others from day 3 – 10 (3 d – 10 d). Patient 2 is infected by patient 1 on day x and risks hospitalization between day x + 7 and x + 13. The arrows indicate expected onset of symptoms on day 5 after infection.

The negative impact of a positive test on hospitalizations of patient 2 relies on that patient 1 was tested positive three to ten days prior (day 3-x to 10-x) and quarantined to avoid infection of patient 2. Hence, if patient 1 is quarantined due to a positive test, it is expected to decrease the number of hospitalizations 7 – 20 days later. This should lead to a negative correlation between the number of tests and the number of hospitalizations. However, no clear correlation was observed during the period of mass testing in Denmark (May 27^th^ 2020 – June 21^st^ 2021) or during each of the mass testing stages 1, 2, and 3 (Table 1). The lack of correlation was independent of the expected delay time (7, 14 or 20 days) between test and hospitalization.

**Table 1.**
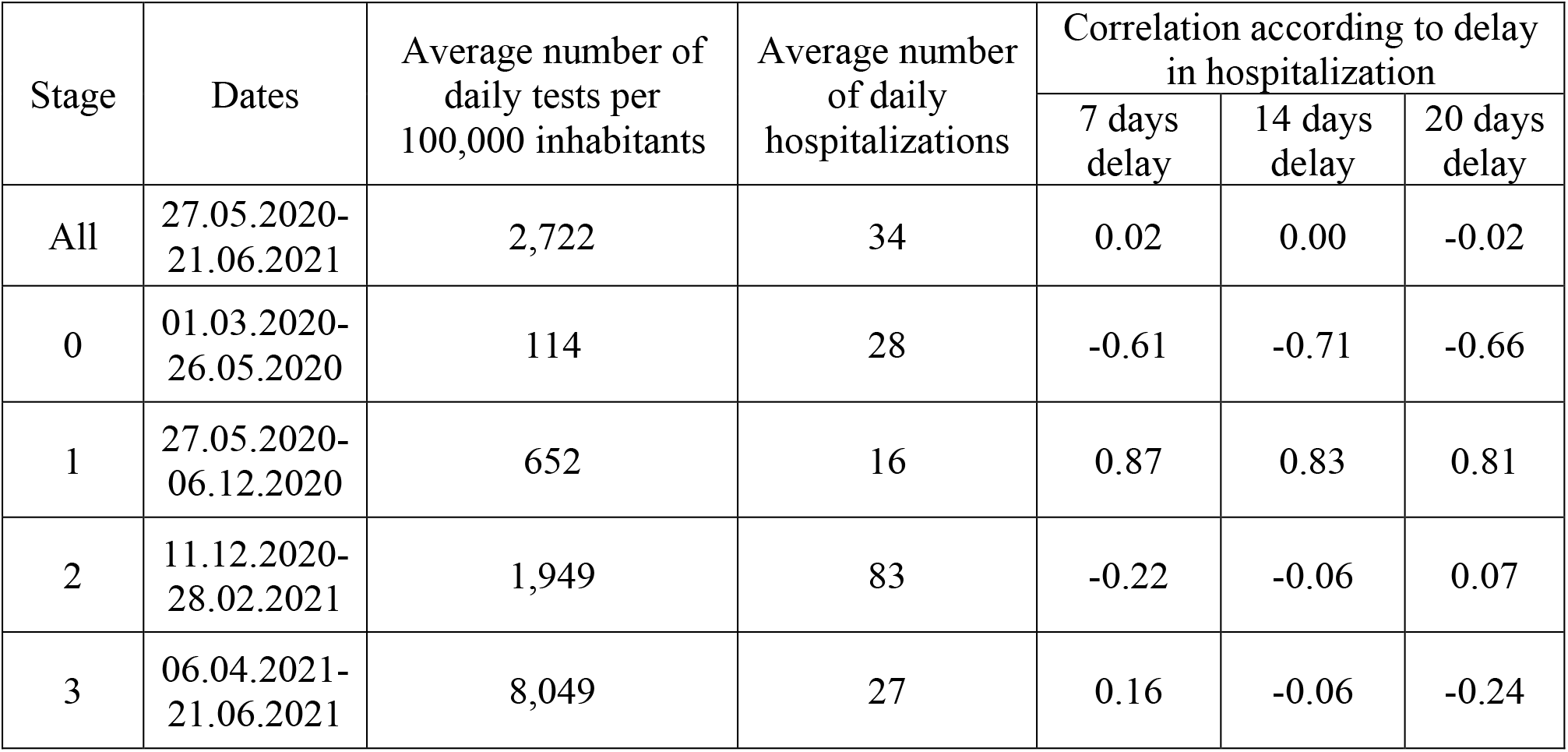
Correlation between number of tests and hospitalizations.

In stage 0 before mass testing was initiated there was a strong negative correlation between the number of tests performed and hospitalizations (Table 1). However, it is not clear whether the decrease in number of hospitalizations during this stage is due to the low-level testing or caused by the lockdown, seasonal variations, or other factors.

Surprisingly, there is a strong, positive correlation between the number of tests and hospitalizations for stage 1 (Table 1). It is obviously that testing per se does not increase contamination with Covid-19 and that the correlation must be due to other factors such as increased awareness of the prevalence of the disease leading to an increased motivation for voluntary testing.

In a voluntary mass testing regime as employed in Denmark, the percentage of the Covid-19 tests that are positive depends on the prevalence of Covid-19 in the population, the number of persons tested, and on alertness in the population. When the number of tests is relatively constant over a shorter period it is reasonable to assume that percentage of positive tests reflects the prevalence of Covid-19 in the population. To further assess the effect of the mass testing in Denmark during stage 3 the positive percentage was calculated for all days in the period April 6^th^ to June 21^st^ 2021 where the total number of tests was between 444,500 and 512,313 (Figure 3). Although the number of tests was almost constant (477,215 tests ± 7 %) the percentage of positive tests increased slightly from 0.22 on April 12^th^ to 0.31 on May 26^th^ and then decreased rapidly to 0.07 on June 17^th^ (Figure 3). In the week before April 12^th^ the average percentage positive tests was 0.24 on an average of 375,000 tests per day whereas in the week after May 26^th^ the average percentage positive tests was 0.27 on an average of more than 525,000 tests per day.

**Figure 3.**
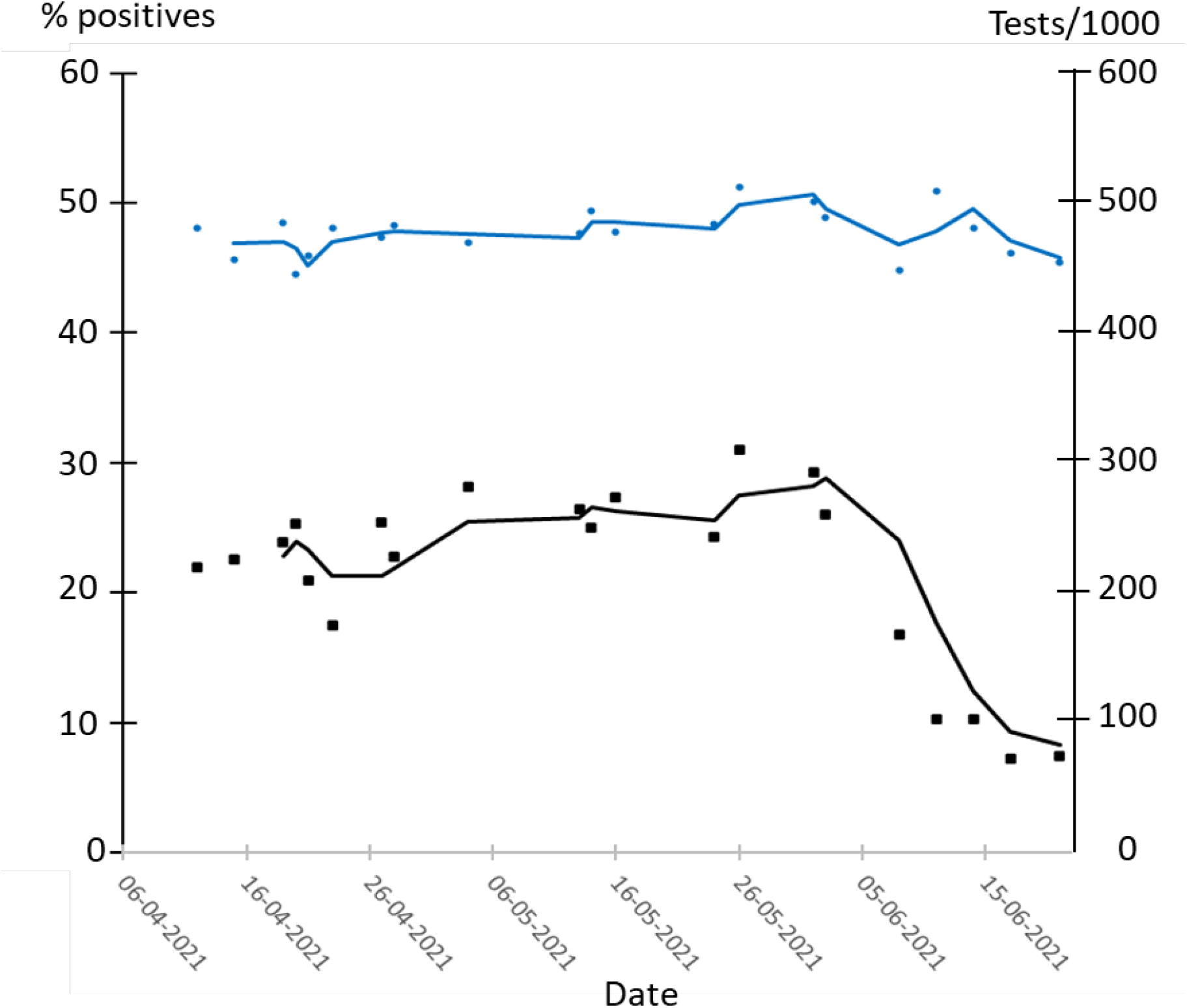
Percent positive tests at approximately the same number of daily tests during April – June 2021. Blue dots: Number of tests. Blue line: Three days average of number of tests. Black dots: % positive tests. Black line: Three days average of % positive tests.

Testing is expected to reduce the number of patients testing positive within x+3 days to x+10 days (Figure 2). Hence, for x between 3 and 10 mass testing should reduce the percentage of positive tests within 6 – 10 days. If the percentage of positive tests reflects the prevalence of Covid-19 the increase in percentage positives over the 44 days of mass testing between April 12^th^ and May 26^th^ does not indicate that mass testing effectively reduced the prevalence of Covid-19 in the Danish population. The decrease in percentage positives below 0.22 after June 2^nd^ (Figure 3) came too late to be a direct effect of the mass testing and may be a seasonal effect or an effect of vaccination of more than half of the population (Danish Health Authority, 2021; Engelbrecht and Scholes, 2021; Ritchie et al., 2020).

Sensitivity of the antigen tests relative to the rt-PCR test is on average 78 % during the first week after symptom onset decreasing to 51 % during the second week (Dinnes et al., 2021). In patients later than the second week after symptom onset the sensitivity drops even further. Hence, the percentage of PCR positive tests that are also positive in antigen test can be used to evaluate whether the tests are performed early in the infection cycle (week 1) when the patient is able to infect others (Figure 2). Curiously, for the period from February 1^st^ to June 20^th^ 2021 the antigen tests only detect Covid-19 in 48 % of the samples that are positive in rt-PCR. This is much lower than the expected 78 % if the patients were tested in week 1 after symptom onset (Dinnes et al., 2021). The relation between positive antigen tests and positive rt-PCR tests for the same patient suggests that many patients detected positive by rt-PCR may be in week 2 or later where they are no longer highly infectious.

To be effective for reducing Covid-19 prevalence and hospitalizations, mass testing should detect infected patients before symptom onset or early in week 1 after symptoms develop. Hence, as testing was increased in Denmark during Spring 2021 the percent of rt-PCR positives that are also detected by antigen tests should also increase. Antigen tests are most effective in the beginning of the infection. However, from February 2021 to May 2021 when testing peaked the percent of rt-PCR positives that were also detected by antigen tests decreased from 60 to 44 % before increasing to 53 % in the first three weeks of June (Figure 4). The average ratio of antigen positives to rt-PCR positives on the same patients for February – March was 48 % whereas testing of the target group of patients in week 1 of the Covid-19 infection should yield a ratio of 80 % or more (Dinnes et al., 2021). Assuming a sensitivity of the antigen test of 0.8 in week 1 and 0.3 in weeks 2 and later the observed ratio between antigen positives and rt-PCR positives suggests that only 36 % of the tested persons are in the target group for testing (0.48 = (0.8 x w1 + 0.3 x w2)/(w1 + w2), where w1 is PCR positives in week 1, w2 is PCR positives in week 2 and w1 + w2 = 100). However, this number should be regarded as a lower limit as it depends on several assumptions about the testing.

**Figure 4.**
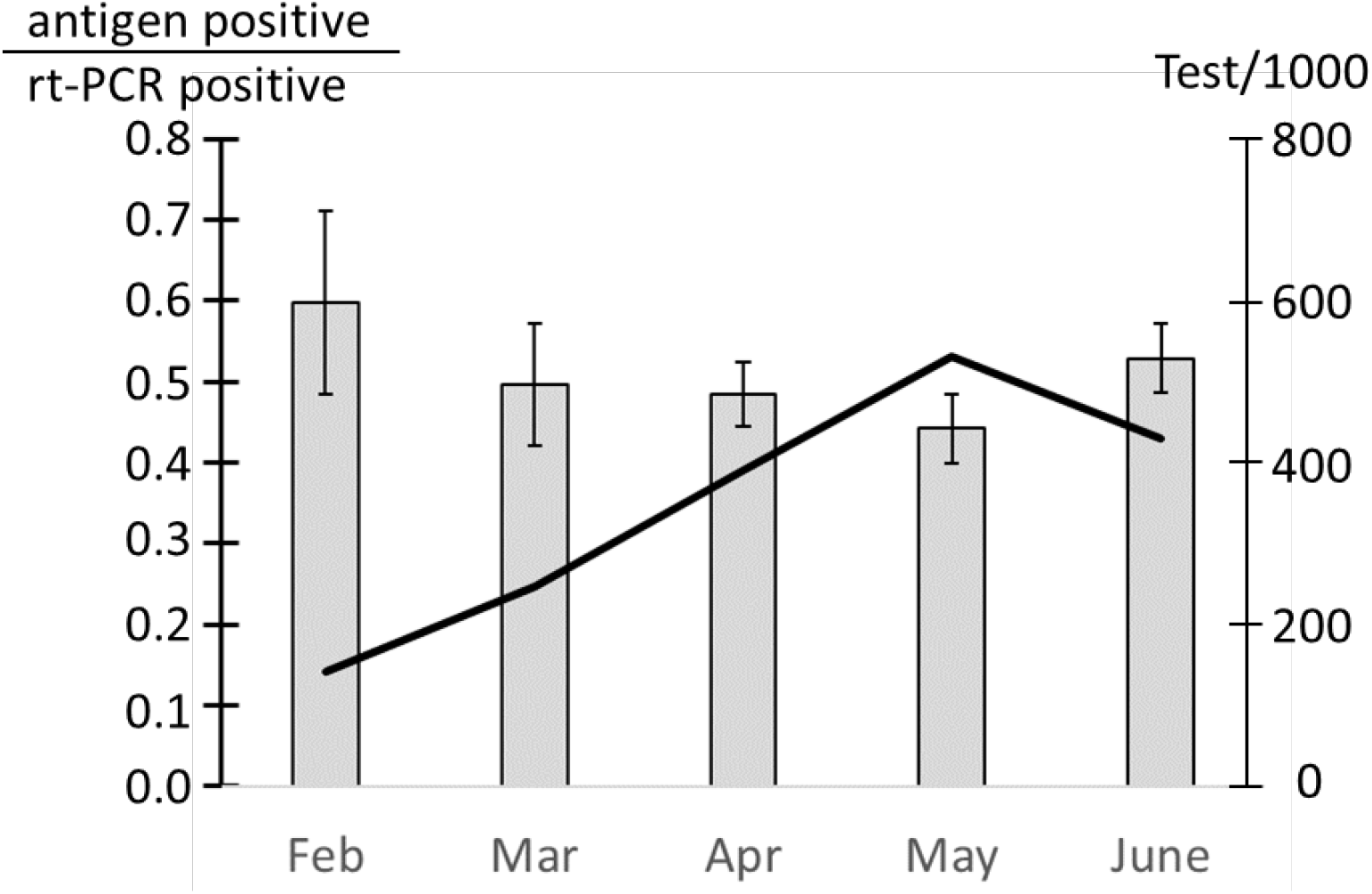
Variation in the number of positive rt-PCR samples that are positive in an antigen test in the Spring 2021. Grey bars: Fraction of rt-PCR positive test that are also positive in an antigen test ± standard deviation. Solid line: Total number of tests performed

It is of assumed that antigen test and PCR test are performed at the same time. This is not necessarily the case. The numbers of positives are based on antigen and rt-PCR tests performed within 48 hours. However, if the PCR test is performed later than the antigen test, patients in the first week of infection may have had time to increase their viral load and thus be more likely to test positive. This will skew the ratio towards lower numbers. Nevertheless, even a sensitivity of the antigen tests as low as 0.6 during the first week of infection implies that 40 % of the rt-PCR positives are outside the target group for testing.

In contrast to what was observed for testing, there was negative correlation of -0.50 between the accumulated number of vaccinations and hospitalizations 14 days later for the period when vaccination was initiated on December 27^th^ 2020 to June 21^st^ 2021 (Figure 5). For stage 3 (April 6^th^ – June 21^st^ 2021) the correlation was -0.66 in agreement with a significant effect of vaccinations on the number of hospitalizations.

**Figure 5.**
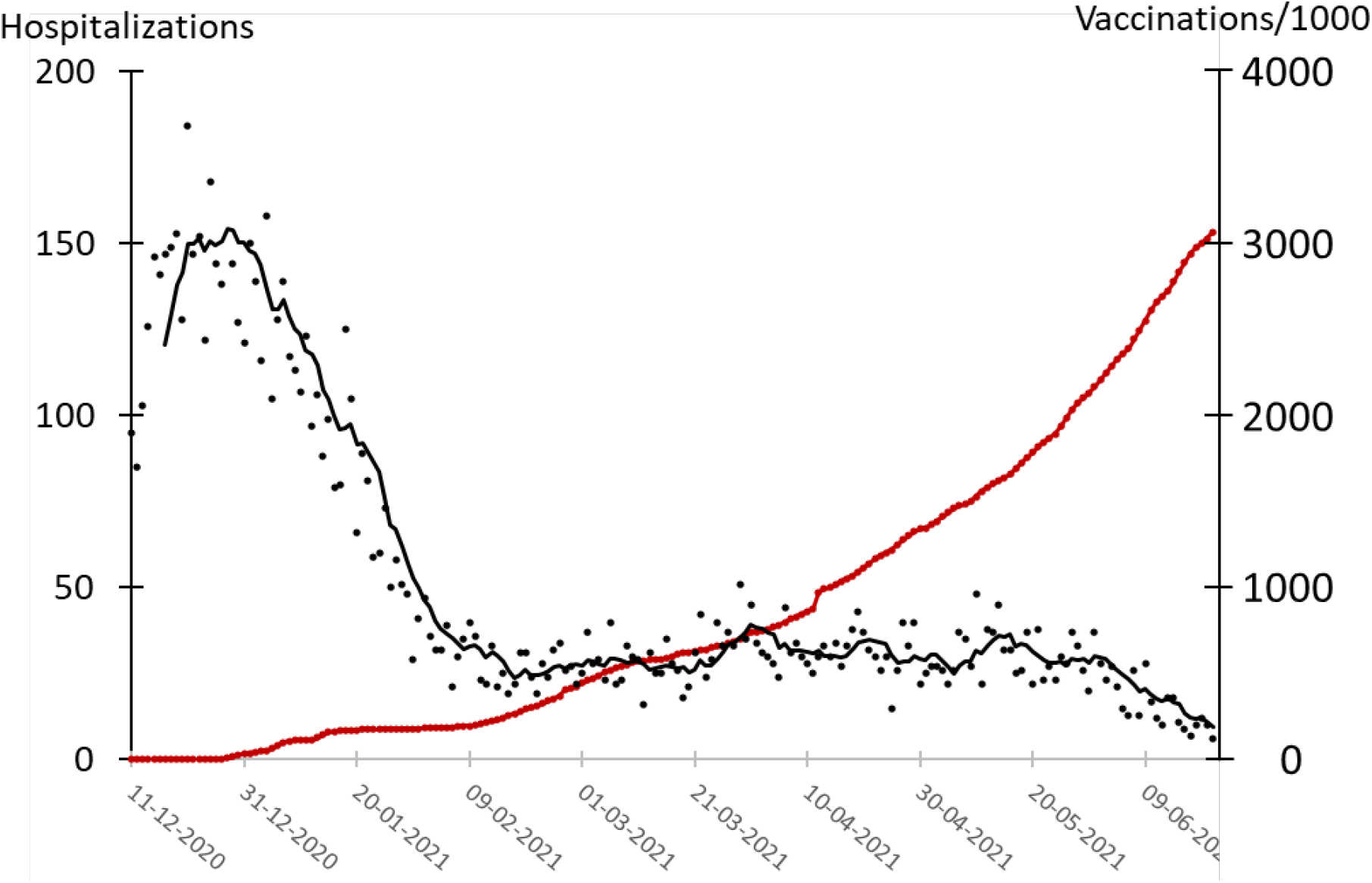
Number of hospitalizations and vaccinations in Denmark during the pandemic. Black dots: Hospitalizations. Black line: Seven days average of number of hospitalizations. Red line: Accumulated number of vaccinations.

## Discussion

The present investigation did not find evidence to support that mass testing effectively reduces the prevalence of Covid-19 in Denmark, as the number of tests and hospitalizations did not correlate. Especially, the lack of correlation during stage 3 from April 6^th^ to June 21^st^, 2021, when an average of more than 8,000 persons per 100,000 inhabitants were tested per day, does not support mass testing as implemented in Denmark, as a tool to reduce prevalence of Covid-19. Testing of this magnitude is sufficient for testing the entire population in less than two weeks in a scheme resembling the mass testing that took place in Slovakia in the Fall 2020 (Pavelka et al., 2021). The systematic testing in Slovakia reduced Covid-19 prevalence by 70 % compared to unmitigated growth and by 58 % in absolute numbers within a week of testing (Pavelka et al., 2021).

Although it cannot be excluded that mass testing in Denmark during Spring 2021 impaired an increase in the number of hospitalizations and the percentage of positive tests, there is no evidence to support this hypothesis. This is in contrast to vaccinations against Covid-19 where the present analysis found the expected correlation between the accumulated number of vaccinations and hospitalizations in agreement with that vaccination protects against hospitalization due to Covid-19 (Ledford, 2021). If the Danish test strategy had had a similar impact as testing in Slovakia in the Fall 2020, it would have shown up as a significant negative correlation between testing and hospitalizations and a decrease in the percentage of positive tests. However, there was no significant rapid decrease in the percentage of positive tests when comparing the percentage of positives on days with similar numbers of tests during stage 3. It is reasonable to attribute the decrease in the percentage of positives towards the end of stage 3 to an improvement in the weather conditions and vaccination of more than one third of the population by the end of May. Both Summer weather conditions and vaccinations reduce the prevalence of Covid-19 (Engelbrecht and Scholes, 2021; Haas et al., 2021).

In Slovakia, it was observed that the effect of the massive testing was limited to regions with high Covid-19 prevalence whereas testing did not seem to have an effect in regions with low prevalence (Pavelka et al., 2021). This lead to the suggestion that a high prevalence, which was not the case in Denmark during Spring 2021, is a prerequisite for any impact of mass testing (Mercer and Salit, 2021).

Another difference compared to Slovakia is that Denmark has not performed a systematic testing of all citizens. As a consequence, testing is unequally distributed reflected in the fact that 10 % of the population aged more than five years has not been tested for Covid-19 (Statens Serum Institut, 2021a). Unequal testing implies that some people are tested more often than appropriate whereas other are tested sporadically or not at all. This distribution of testing is likely to be involved in lowering the ratio between rt-PCR positives and antigen test positives. The most plausible explanation for this low ratio is that a large fraction of the testing is not performed on the target group of infectious persons but on people with late-stage Covid-19 where the risk of transmission is low or absent. In a strict sense, PCR positives with late-stage Covid-19 can be regarded as false positives. The estimation that only 36 % of the PCR positives are in the target group for testing implies that 64 % of the PCR positives are false. This number should be regarded as an upper limit as it depends on a number of assumptions about the testing. Nevertheless, the results suggest that a large fraction of the PCR positives are a kind of false positives as these persons are likely to have late-stage Covid-19 with low or absent risk of infecting others.

An implication of such a large fraction of Covid-19 infections passing undetected until late-stages is that the mass testing is not efficient in finding the patients that are infectious.

From a theoretical point of view, it is logical to assume that detection of Covid-19 positives by mass testing in order to quarantine infected individuals and break chains of infection reduces prevalence of the disease. A recent bottom-up calculation reached the conclusion that the Danish mass testing in the Spring 2021 reduces the prevalence of Covid-19 by removing 25 % of the infected patients every day (Pedersen et al., 2021). This conclusion is based on an extrapolation of the expected number of positive tests if Denmark had followed a low-testing strategy like the neighbouring countries Norway and Sweden (Pedersen et al., 2021). The extrapolation assumes that Denmark would have performed 30,000 tests per day leading to the diagnostics of approximately 200 Covid-19 positive patients. The authors express their reservations as to whether the extrapolation is valid under the conditions in the Spring 2021. Indeed, the actual testing carried out in Norway in the same period detected on average 550 Covid-19 positives per day although only 16,000 – 27,000 tests were performed (Ritchie et al., 2020). Hence, the assumption that 30,000 tests per day would only detect 200 positives (Pedersen et al., 2021) seems to be a large underestimation. On the prerequisite that the Danish health authorities are as efficient as the Norwegian on Covid-19 testing it is safe to conclude that low-level testing in Denmark would have identified more than 550 Covid-19 cases per day in the Spring 2021. If the bottom-up calculations (Pedersen et al., 2021) are adjusted to this notion, the impact of mass testing during Spring 2021 was to remove not more than 10 % of the infected persons per day.

In this context it is interesting that there was a correlation between the number of tests and reduction of hospitalizations in Denmark during the Spring 2020 when testing was performed at a low level. This correlation raises the possibility that low-level testing of symptomatic patients and close contacts has a positive impact on control of the pandemic but that further increase in testing is futile. However, the observed, negative correlation should be interpreted carefully as other measures such as an increased awareness of the pandemic, lockdown and weather conditions are likely to have had a positive effect on the number of hospitalizations in the Spring 2020 (Engelbrecht and Scholes, 2021; Jun et al., 2021).

It is reasonable to assume that such a low impact of mass testing (10 % decrease in prevalence) would not be detectable when comparing the number of tests to hospitalizations. Hence, a bottom-up calculation based on the assumption that the Danish health authorities are as efficient as the Norwegian rather than on a theoretical extrapolation from large-scale to low numbers would agree with the observations in the present study that mass testing does not have a measurable impact on Covid-19 prevalence in Denmark.

The question remains as to why mass-testing does not impact Covid-19 prevalence. One reason could be, as described in the present study, that the testing for a large part is performed outside the target group of early-stage infectious persons. This could be attributed to the unsystematic approach to testing in Denmark.

Lieberman, Lieberman and Bourassa suggested that testing induces unsafe behaviour leading to increased transmission of Covid-19 thereby outweighing the positive effects of testing (Bourassa, 2020). According to this possibility, individuals tested negative ignores the considerable risk of false negatives of 0.20 and 0.15 respectively for the antigen and the rt-PCR tests (Dinnes et al., 2021; Hu et al., 2020). Instead, the negative test result increases the risk behaviour. This is a general phenomenon in diagnostics described as preventive misconception (Simon et al., 2007). A simple thought experiment can demonstrate how elevated risk behaviour can outweigh mass testing: The risk of transmission of Covid-19 from one person to another depends on the third power of the distance between the two. If mass testing removes 25 % of the Covid-19 infected patients every day as suggested (Pedersen et al., 2021) it only takes a 10 % decrease in the average distance between the rest of the citizens to offset the effect on airborne transmission. Thus, only a small increase in risk behaviour regarding distance and other Covid-19-related precautions can neutralize the effect of mass testing. It seems that preventive misconception leading to increased risk behaviour is a plausible explanation for the missing impact of mass testing.

## Conclusion

The Danish mass-testing strategy for Covid-19 was found not have an effective impact on the number of hospitalizations due to Covid-19. Furthermore, the ramp up of the mass testing during the Spring 2021 did not lower the number of positive tests at comparable test numbers. On the contrary, the number of positive tests increased slightly between April 12^th^ and May 26^th^ thus underpinning that mass testing did not reduce the prevalence of Covid-19, at best.

A likely explanation for the lack of impact of mass testing is that the testing increases the risk behaviour of the tested persons, and that the Danish testing was done in a very unfocused manner. Thus, the high fraction of false negatives at low Covid-19 prevalence combined with the increased the risk behaviour fully outweigh the beneficial effects of mass testing.

## Data Availability

All data are available.

## Acknowledgements

None.

